# Feasibility Study of the CT Clock Tool for Estimating Onset Time of Ischaemic Stroke: Study Protocol

**DOI:** 10.1101/2023.09.08.23295203

**Authors:** Grant Mair, William Whiteley, Francesca Chappell, Allan MacRaild, Joanna Wardlaw

**Affiliations:** Centre for Clinical Brain Sciences, University of Edinburgh, UK; Royal Infirmary of Edinburgh, NHS Lothian, Edinburgh, UK

## Abstract

Ischaemic stroke is a devastating disease with high rates of death and disability affecting ∼100,000 people annually in the UK. Effective treatments are only offered when the time of stroke onset is known and within specific limits. For the 20% where onset time is unknown or delayed, advanced imaging methods can identify people for safe and effective treatment. However, this advanced imaging is not widely available. We have developed a method for identifying patients who can still be treated even without advanced imaging. The CT Clock Tool uses only the non-enhanced CT brain scan, that most patients with stroke receive upon arrival at hospital, to determine whether patients are eligible for treatment.

This project aims to provide the first prospective clinical testing of the CT Clock Tool in a single-centre feasibility analysis. We will recruit patients in the emergency department with ischaemic stroke and obtain consent to use their acute CT brain imaging and other data relating to their care. We will ask treating clinicians to apply our CT Clock Tool method in real time, but we will not alter routine care pathways or otherwise involve patients. We have powered the study to validate previous estimates of the diagnostic accuracy and precision of our method for identifying treatment eligible patients. Results from this project will enable a future definitive randomised-controlled trial, which if successful would allow the routine use of our method anywhere CT brain imaging is available for patients with stroke.

## 1 INTRODUCTION

### 1.1 BACKGROUND

Stroke is a major health concern worldwide causing high rates of disability and death. Most strokes (approximately 85%) are cause by ischaemia following blockage to an arterial blood vessel. Two effective treatments are available that can remove the vessel blockage and thus reduce the rates of both disability and death after stroke. However, both treatments have time-limits for safe delivery. Thrombolysis uses an injected medicine to break down the blood clot and must be used within 4.5 hours of stroke onset (the European licensing time limit for alteplase). Thrombectomy is a surgical procedure that involves pulling the clot out using a catheter placed inside the artery and is usually limited to 6 hours from stroke onset.

Unfortunately, for approximately 20% of patients with ischaemic stroke, the time of symptom onset is not known, usually because they wake with symptoms or are found collapsed.^1^ Such patients have traditionally been denied treatment, but recent research has shown that advanced brain imaging methods (for example CT perfusion or MRI) can be used to identify patients who can still safely be treated in the absence of a known onset time for stroke.^2-5^ However, these advanced imaging methods are not widely available for acute stroke assessment, especially outside major treatment centres in developed nations. For example, with our distributed population in Scotland, it remains difficult to offer advanced brain imaging to all who might need it. Plain (non-enhanced) CT is much more widely available and is the standard immediate assessment method for most patients with stroke worldwide.

As ischaemic stroke progresses, injured brain slowly begins to change on plain CT and the ischaemic lesion becomes more and more pronounced with greater oedema and swelling. These changes are seen as a darkening of tissue, indicating a decrease in CT attenuation, Figure 1.

**Figure 1.**
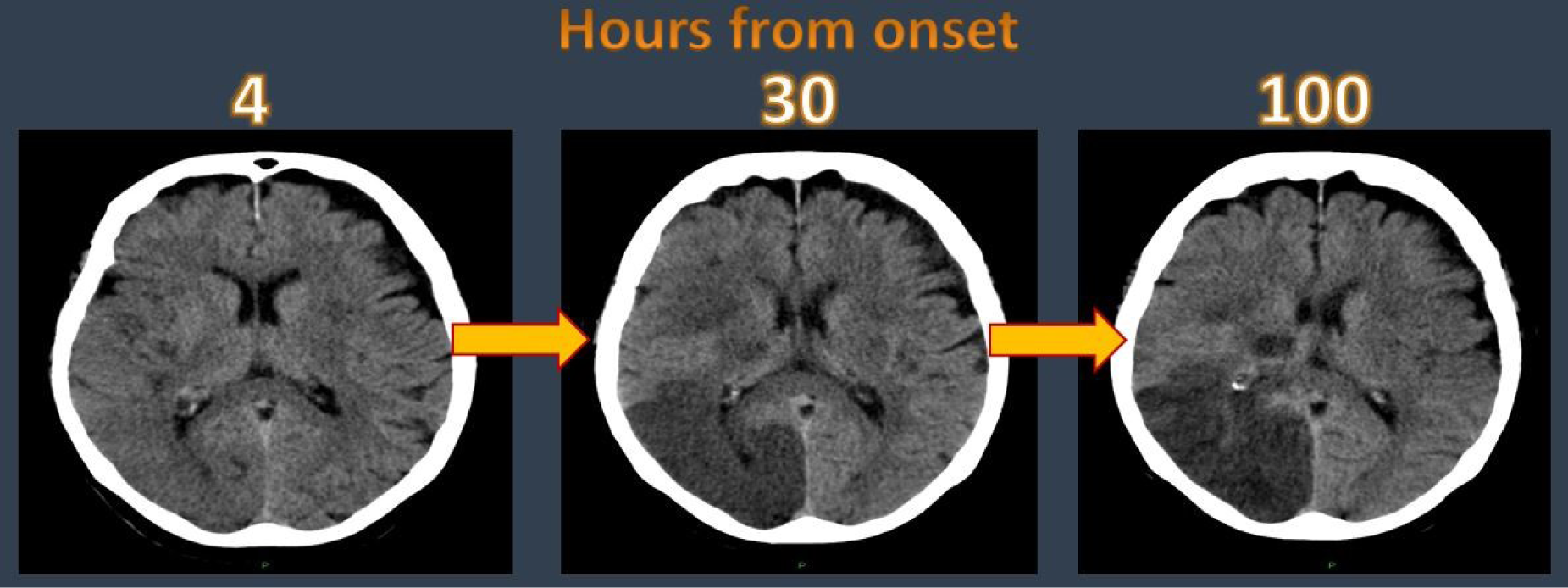
CT attenuation changes of ischaemic brain tissue with time.

We have developed a method for estimating time of ischaemic stroke onset for individual patients that uses only attenuation measurements of plain CT brain imaging, the CT Clock Tool.

### 1.2 RATIONALE FOR STUDY

Our proposed CT Clock Tool method for estimating the time of stroke onset (and thus clarifying individual patient eligibility for effective treatment) is significantly simpler to use in the acute setting than the advanced methods currently available (CT perfusion or MRI).

The CT Clock Tool can provide specific time estimates for a given patient. These estimates are derived from simple measurements of brain that can be manually acquired from non-enhanced CT scans within seconds, no other imaging or specialist viewing software is required. Specifically, users (for example stroke clinicians or radiologists) measure the attenuation of any visible ischaemic lesion and of the equivalent normal brain to derive a CT attenuation ratio (attenuation of ischaemic brain ÷ attenuation of normal brain), Figure 2.

**Figure 2.**
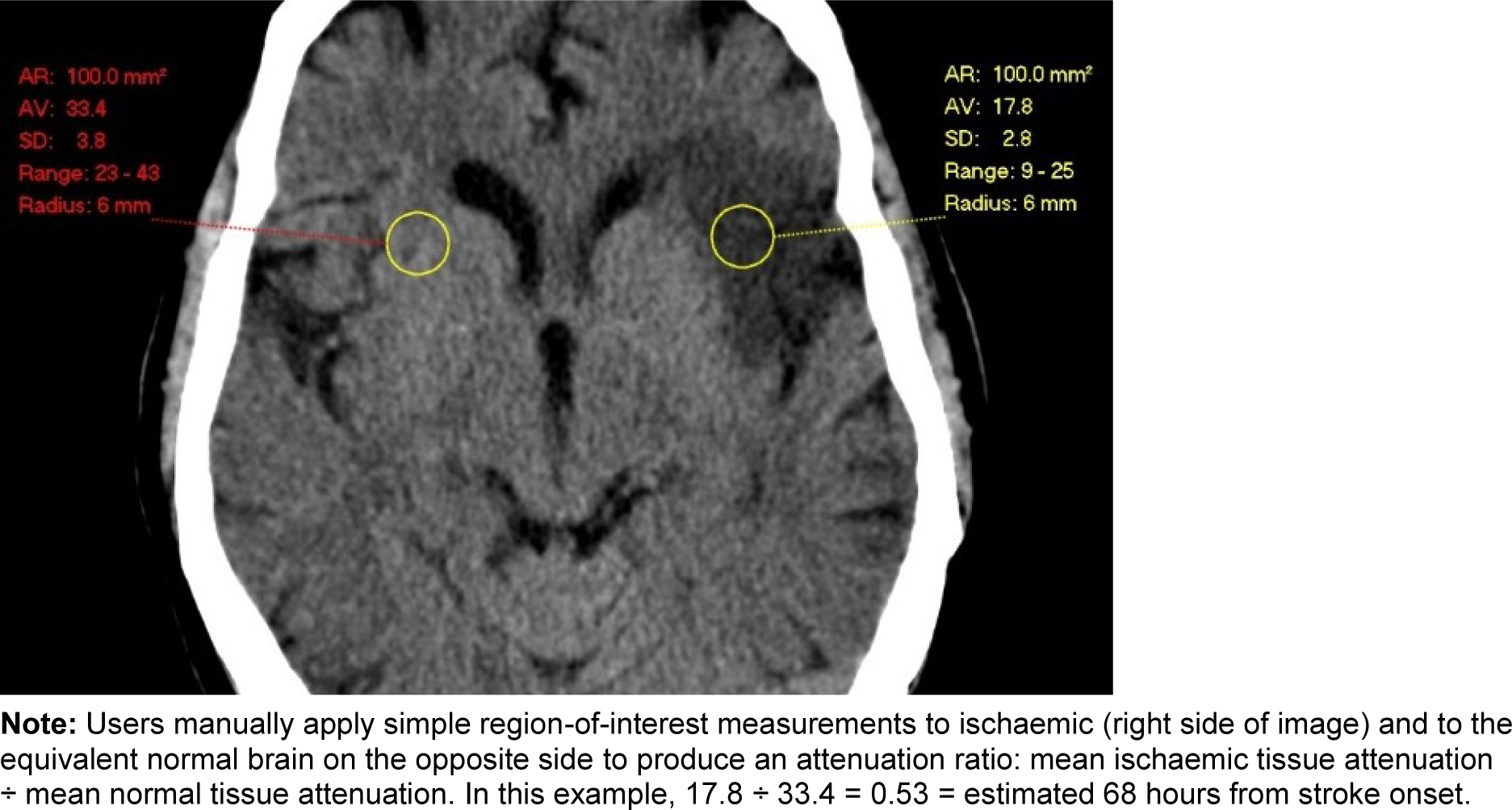
CT Clock Tool method.

Initial testing shows that an attenuation ratio greater than 0.82 is highly predictive of a stroke onset to CT time less than 4.5 hours (i.e. within the time limit for thrombolysis with alteplase). At this threshold, the CT Clock Tool is 97% sensitive and 83% specific for correctly classifying patients who are within 4.5 hours of stroke symptom onset.^6^ In addition, we have tested associations between the appearances of non-enhanced CT and CT perfusion (CTP) when both are acquired at the same time. We compared the CT attenuation ratio with CTP appearances that indicate affected brain tissue is likely to remain viable. In other words, and in the absence of CTP, a CT attenuation ratio greater than 0.87 is 86% sensitive and 91% specific for identifying brain tissue that remains viable (and is therefore most likely to benefit from treatment).^7^ For patients who present to hospital very early after stroke symptom onset, for example within 1 hour, the ischaemic lesion may not yet be visible on plain CT, even to experts (i.e. the attenuation ratio is near normal, very close to 1.0). Therefore, we have also developed a complementary method of identifying patients who are likely to have large ischaemic lesions regardless of whether they can be seen on plain CT; essentially when there is a clinical-radiological mismatch (National Institutes of Health Stroke Scale, NIHSS >11 but a normal CT).^8^ Finally, a recent randomised-controlled trial and two previous small studies have found it is safe to treat patients with thrombolysis in the absence of a known stroke onset time, so long as there are no/minimal obvious ischaemic lesions on their non-enhanced CT scans.^9-11^ However, the randomised trial did not find that thrombolysis was associated with better outcomes at 90 days after stroke.^11^ Combining all these different results, our CT Clock Tool aims to provide a useable output (guidance for the appropriateness of treatment based on an estimate of stroke onset time and injured brain tissue viability) for the greatest proportion of patients with ischaemic stroke but an unknown time of onset.

We have a longer-term aim of incorporating the CT Clock Tool into routine clinical care, especially in centres that cannot provide alternative or advanced methods to assess patient eligibility for acute treatment. The current proposal seeks to provide a first prospective clinical feasibility evaluation to assess the practicalities of using the method in real time and to evaluate the potential impact on clinical care.

We anticipate the results of the present analysis will inform the design of a future randomised-controlled trial to evaluate whether the CT Clock Tool can safely be used to determine the eligibility of patients for treatment with thrombolysis and/or thrombectomy where time of stroke onset is unknown or is delayed.

## 2 STUDY OBJECTIVES

### 2.1 OBJECTIVES

#### 2.1.1 Primary Objective

- Feasibility of front-line clinicians (stroke clinicians and radiologists) using the CT Clock Tool method during routine care.

#### 2.1.2 Secondary Objectives

- Comparison of stroke onset time estimates provided by front-line clinicians using the CT Clock Tool with actual elapsed time.
- Diagnostic accuracy of these time estimates for determining treatment eligibility relative to standard thrombolysis and thrombectomy limits (4.5 and 6 hours, respectively).

○ Whether users should be encouraged to estimate location of ischaemic lesions.
○ Impact of stroke severity on accuracy of estimates.
- Whether estimates of brain tissue viability correlate with equivalent CTP findings.

### 2.2 ENDPOINTS

#### 2.2.1 Primary Endpoint

- Proportion of cases where a front-line clinician successfully uses the CT Clock Tool method in the acute setting, i.e. acquires CT measurements (regardless of result)

#### 2.2.2 Secondary Endpoints

- Accuracy of stroke onset time estimates – absolute error of time estimates, proportion over versus under calling time.
- Sensitivity, specificity, positive and negative predictive values for the CT Clock Tool with reference to thrombolysis and thrombectomy time limits.

○ Compare in sensitivity analyses results for those estimating lesion location and with differing stroke severity.
- Proportion of cases where tissue viability estimates of the CT Clock Tool agree with CTP indicators for tissue viability.
- User experience of the CT Clock Tool, ease of use for finding/estimating the location of lesions and making measurements.
- Inter-rater agreement for CT attenuation measurements and the respective time and tissue viability estimates derived from different front-line clinician measurements of the same CT.

## 3 STUDY DESIGN

Prospective single centre cross sectional 12-month feasibility study.

Study will be based in the Royal Infirmary of Edinburgh (RIE) a comprehensive stroke centre offering thrombolysis, thrombectomy and hyper-acute stroke unit (HASU) care. All patients presenting to RIE with symptoms of stroke are imaged with non-enhanced CT soon after arrival. Thrombolysis is available 24/7. Where patients may be a candidate for thrombectomy, advanced stroke imaging including CT angiography (CTA) and CTP are also provided at baseline. Thrombectomy (and thus, advanced stroke imaging) is currently only available weekdays 08.00-20.00.

Patients will be identified and recruited from the RIE emergency department with assistance from the EMERGE (Emergency Medicine Research Group of Edinburgh) nurses. Application and testing of the CT Clock Tool will occur during the patient’s presentation to the RIE emergency department by the clinicians involved in that patient’s care. Specifically, we will invite the treating stroke clinicians and radiologists reviewing the stroke CT imaging to take part (prior to the study start date, treating clinicians will be alerted to the study plan and offered training in the CT Clock Tool method). To avoid delaying emergency care, we will encourage clinicians to complete testing once all urgent tasks are complete (we suggest stroke clinicians complete the Clock Tool assessment when writing their notes, and radiologists after they have completed their regular CT report). There will be no direct involvement of patients (other than recruitment) and no anticipated alteration to their routine care. To maximise the chance of finding an ischaemic lesion on brain CT that is big enough to measure, we will not recruit patients with minor or lacunar stroke.

Finally, we will conduct a delayed (approximately 2 weeks after recruitment) review of each participant’s medical record, to confirm that final diagnosis was ischaemic stroke and to complete data collection. Again, there will be no direct patient contact during this phase.

Treating clinicians are also considered as research participants in this study. Treating clinicians will be approached prior to the start of patient recruitment and invited to take part. Treating clinicians will be offered training in the CT Clock Tool method.

## 4 STUDY POPULATION

We will recruit adult patients presenting acutely to the RIE emergency department with symptoms of stroke, where non-enhanced brain CT excludes haemorrhage and other structural causes for symptoms (i.e. ischaemic stroke is the most likely diagnosis).

We will also recruit front-line clinicians who routinely deliver care to our target patient population. Specifically, those clinicians involved in making treatment decisions for patients with ischaemic stroke. This includes stroke clinicians, radiologists, and neuro-interventionalists.

### 4.1 NUMBER OF PARTICIPANTS

To confirm whether our previous estimates for CT Clock Tool sensitivity and specificity are sufficiently precise, we estimate that 118 CT scan assessments are required. See section 9.1 for detailed patient sample size calculation.

All clinicians who routinely review the CT brain imaging of patients with acute ischaemic stroke at RIE will be invited to take part. We do not set a minimum number for clinician participants, but at any given time there are estimated to be over 60 such clinicians. This includes approximately 20 stroke clinicians, 35 radiologists and 8 neurointerventionists. We anticipate that at least 50%, or around 30 clinicians will agree to take part.

### 4.2 INCLUSION & EXCLUSION CRITERIA

#### 4.2.1 Inclusion Criteria for Patients

- With and without a known time of stroke onset.
- With a stroke severity score (NIHSS) >4
- With suspected anterior or posterior circulation ischaemic stroke.
- Over 18 years of age with no upper limit.
- Patients with and without concurrently acquired CTP imaging (acquired only for potential thrombectomy candidates, approximately 20%)
- Patients able to provide informed consent (including witnessed consent when loss of functional ability is evident, e.g. upper limb weakness in dominant hand or visual loss).

#### 4.2.2 Exclusion Criteria for Patients

- With a suspected lacunar syndrome (i.e. pure motor stroke, pure sensory stroke, mixed sensorimotor stroke [without cortical signs], ataxic hemiparesis, dysarthria-clumsy hand syndrome).
- Where CT brain imaging demonstrates a non-ischaemic cause for stroke symptoms, e.g. brain haemorrhage, tumour or other relevant structural abnormality.

#### 4.2.3 Inclusion Criteria for Clinicians

- Who routinely review CT brain imaging of patients presenting with acute stroke symptoms to the Emergency Department at RIE.
- Who are actively involved in making treatment decisions for patients with ischaemic stroke.
- Stroke specialists, radiologists and neurointerventionists.
- Consultant and training grade clinicians.
- Over 18 years of age with no age limit.
- Able and willing to receive training in the CT Clock Tool method.

#### 4.2.4 Exclusion Criteria for Clinicians

- Clinicians who are not physicians
- Emergency department clinicians.

### 4.3 CO-ENROLMENT

Excepting the time required for recruitment and for clinicians to assess imaging, our study should have no direct impact on routine care. Thus, there is no specific barrier to co-enrolment, regardless of other trial type.

Co-enrolment will be handled by the EMERGE team, who will assess individual patient/family suitability for co-enrolment with respect to the anticipated burden for participants and in line with the Sponsors co-enrolment policy.

There are no restrictions on clinicians to be co-enrolled in other studies, but this is deemed unlikely.

## 5 PARTICIPANT SELECTION AND ENROLMENT

### 5.1 IDENTIFYING PARTICIPANTS

#### 5.1.1 Patients

The EMERGE team are active within the RIE emergency department during normal working hours, i.e. Monday-Friday, 08.00-16.00. In close discussion with treating clinicians, the specialist stroke research nurses within EMERGE review patients presenting with stroke as potential candidates for any of the active stroke trials within their portfolio. Following review of individual trial eligibility criteria and discussion with the treating clinicians to confirm eligibility, patients are approached by the EMERGE team regarding any trials for which they may be suitable. The appropriateness of co-enrolment is also considered case-by-case, see section 4.3.

#### 5.1.2 Clinicians

Clinicians who may participate will be identified by role and during routine practice. The Chief Investigator (CI) is also an NHS Lothian radiologist who routinely delivers stroke care. In other words, the CI will invite colleagues to participate (and will offer training) in the weeks prior to starting patient recruitment.

### 5.2 CONSENTING PARTICIPANTS

#### 5.2.1 Patients

Following discussion with the treating stroke clinician, the EMERGE stroke research nurses will approach potential candidates for our study during the period of care within the emergency department. However, given that our study will proceed separate to care, this approach can and will preferably be made once immediate care decisions are complete and hyperacute treatment pathways initiated (e.g. administration of intravenous thrombolysis, referral for thrombectomy, reporting of CT).

Patients will be provided with the study Patient Information Sheet, PIS.

The key messages for potential candidates are:

1. Their decision to be involved or to not be involved in the study will not affect their care.
2. There is nothing for them to do.
3. We only want to use their CT scan and other associated data about them and the care they receive during the current hospital visit to test our CT Clock Tool method.
4. We will retain and share their data for future research, after removing their personal details.

Given the need to test the CT Clock Tool method during routine care, patients will routinely be given approximately 5-10 minutes to consider whether to participate in the study.

Consent will be obtained by a member of the EMERGE team, who will also manage handling and storage of completed consent forms.

Some patients with stroke may not be physically able to sign a consent form. We will therefore include a section for witness certification of verbally given consent.

#### 5.2.2 Clinicians

Potentially participating clinicians will be offered the study Clinician Information Sheet, CIS.

The key messages for potential candidates are:

1. Participation is entirely voluntary.
2. The study budget will refund NHS Lothian the expected cost of their time participating.
3. We will not record personally-identifiable information about them (other than their name on the consent form and to ensure recruited clinicians are appropriately prompted to complete assessments for recruited patients under their care, we will add their name to a list of active clinician participants).
4. Study results will not be incorporated into the NHS record for patients who participate.

Clinicians will be offered as much time as they need to decide whether or not to take part. Where clinicians cannot decide on the same day they are invited, the CI will revisit the question after an agreed time period, e.g. 24 or 72 hours.

Consent will be obtained by the CI, who will also manage handling and storage of completed consent forms.

### 5.3 Withdrawal of Study Participants

Participants are free to withdraw from the study at any point or a participant can be withdrawn by the Investigator (if there is loss of capacity).

If withdrawal occurs, the date and primary reason for withdrawal will be documented in the participant’s case report form (patients) or on their consent form (clinicians), if possible.

The participant will have the option of withdrawal from:

1. All aspects of the study but with continued use of data collected up to that point. To safeguard rights, the minimum personally-identifiable information possible will be retained.
2. All aspects of the study without further use of their data.

Where withdrawal occurs due to loss of capacity, option 1 will apply.

## 6 STUDY ASSESSMENTS

### 6.1 STUDY ASSESSMENTS

EMERGE nurses will assess the eligibility of potential participating patients, acquire consent, and collect screening and baseline data of patients.

CT Clock Tool assessment will be completed by recruited stroke clinicians, radiologists and neuro-interventionists involved in the care of the recruited patients.

The CI will collect follow-up data around 2-3 weeks after patient enrolment using all available clinical and imaging information.

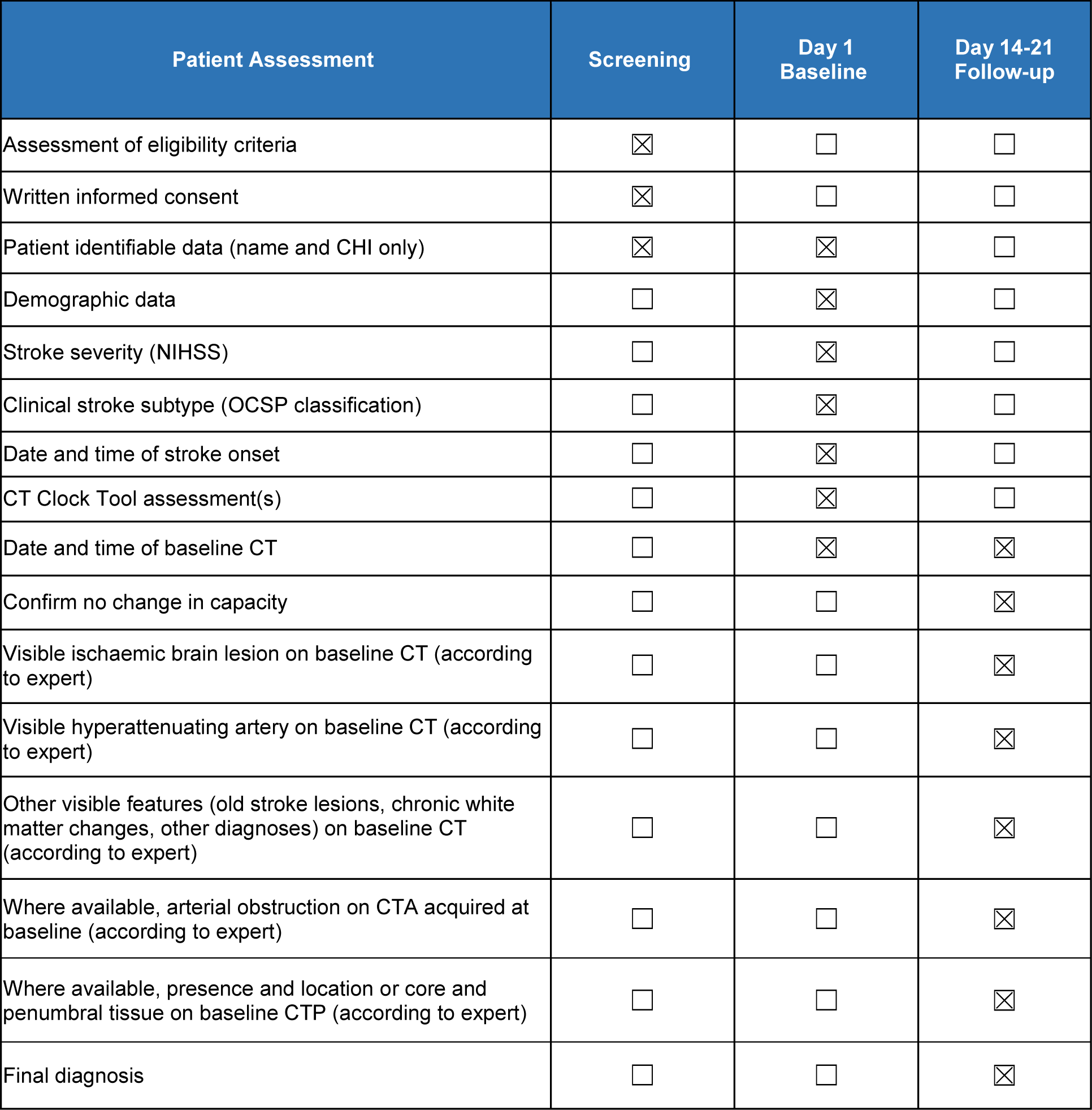

## 7 DATA COLLECTION

### 7.1 Data for Collection

#### 7.1.1 Screening data

Collected by EMERGE nurses and recorded in the paper *Subject Pre-Screening Log* (identifying potential participating patients) and the *Consent & Subject Status Log* (confirmation of consent and details of patients):

- Initials of possible participant
- Date and time of approach
- Initials of nurse making approach and completing screening
- Eligibility criteria met – Y/N
- Consent
  ○ Obtained - Y/N
    ▪ If Y, study number (sequentially applied from CTC001)
    ▪ To ensure study identification numbers are unique with no gaps, the *Pre-Screening Log* provides the only source for study ID allocation
- Reasons for non-recruitment.

**No patient identifiable data will be recorded in the *Subject Pre-Screening Log*.**

**For recruited participants only, the *Consent and Subject Status Log* will include patient identifiable data (name and CHI) in addition to study ID and therefore acts as the study pseudonymisation key.**

Following recruitment of patient participants, EMERGE nurses will:

- Prepare the CT Clock Tool paper reminder forms and the CT Clock Tool paper assessment forms, see 7.1.2.2.
  ○ Preparation of these forms includes adding the participant’s unique study number, date of recruitment, and to ensure data are collected from the correct patient, two forms of patient identifiable data – name and CHI (Community Health Index) number.
  ○ Using the reminder form, EMERGE nurses will then prompt stroke clinicians involved in the case (and who are already recruited to the study) to take part, offering the paper assessment form where required.
- Alert the CI that a patient has been recruited (and if treating stroke clinicians have not yet been invited to participate).
  ○ CI will then prompt radiologists and neuro-interventionists involved in the case (and who are already recruited to the study) to take part (via the relevant radiology departments), also using paper reminder forms.
  ○ CI will invite and attempt to recruit any treating clinicians not yet aware of the study.
- Collect baseline clinical and personal data.

#### 7.1.2 Baseline data

##### 7.1.2.1 Clinical and identifiable patient data

Collected by EMERGE nurses - for practical purposes and to ensure accurate data collection, recorded in a paper CRF and including patient identifiable data:

- Participant name and CHI
- Participant study number
- Confirmation that eligibility criteria have been met– Y/N for each (all Y required)
  ○ Acute stroke symptoms
  ○ ≥18 years old
  ○ Able to provide informed consent
  ○ Brain CT imaging acquired
  ○ Ischaemic stroke is most likely diagnosis (i.e. no haemorrhage or other [non-stroke] abnormality on baseline CT to account for symptoms)
  ○ NIHSS >4
  ○ Not a suspected lacunar stroke syndrome.
- Demographics – age, sex
- Date and time of baseline CT imaging
- Date and time of stroke symptom onset, or last known well, or wake up time – 24-hour clock, to the nearest minute, or unknown.
- NIHSS
- Clinical stroke subtype – OCSP (Oxford Community Stroke Project) classification:
  ○ Total Anterior Circulation Syndrome
  ○ Partial Anterior Circulation Syndrome
  ○ Posterior Circulation Syndrome
  ○ Lacunar Syndrome
- Date & time of form completion

##### 7.1.2.2 CT Clock Tool Assessment(s)

Collected by clinicians and recorded in either the electronic or the paper CT Clock Tool assessment form:

- The choice of electronic or paper form will depend on whether the contributing clinician(s) have access to the internet, electronic is preferred:
  ○ As there is no internet access in RIE A&E resuscitation (where severe strokes are received), limited internet access elsewhere in RIE A&E, but normal internet access in radiology, we will provide both an electronic and a paper form for collecting CT Clock Tool assessment data.
  ○ Both forms include a brief reminder of the study, the participant’s unique study number, and the date and time of their baseline CT.
  ○ The CT Clock Tool reminder form also includes the study web address (www.ctclock.net) and details of generic login details for clinicians, see 8.3.2.
- We will invite stroke clinicians and radiologists (front line clinicians) involved in the routine care of recruited participants to complete the assessment, this may include:
  ○ Consultant stroke clinician
  ○ Specialist trainee stroke clinician
  ○ Consultant radiologist
  ○ Specialist trainee radiologist
  ○ Consultant interventional neuroradiologist
  ○ Specialist trainee interventional neuroradiologist
- At least one clinician from each group, (each clinician must be involved in the participant’s care) will be invited to complete a CT Clock Tool assessment for each recruited participant.
  ○ EMERGE nurses will invite stroke clinicians in the emergency department
  ○ CI will invite radiologists
  ○ Each invited clinician will be handed either a CT Clock Tool reminder form or a CT Clock Tool assessment form and will be asked to complete the CT Clock Tool assessment.
  ○ CT Clock Tool assessments should ideally be conducted in real time, but preferably after immediate patient care is completed. We will accept submissions at any point within the day of recruitment (to avoid clinician bias from non-baseline data, we will reject submissions provided after the day of recruitment).
- We expect therefore, that multiple assessments will occur in parallel for each recruited participant.
- Clinicians will input data derived from the participant’s CT imaging, which should be viewed in parallel with the web-based, or paper form.
- Both electronic and paper CT Clock Tool assessment forms include guidance for using the CT Clock Tool method.

The following user-provided data will be collected via the electronic and paper CT Clock Tool assessment forms (options per question provided in brackets):

- *Section 1* - Patient and clinician details
  ○ Patient participant study number (free text)
  ○ Grade (Consultant/Specialist trainee) and speciality (Stroke/Radiology/Interventional Neuroradiology) of clinician completing assessment.
  ○ Date of assessment
- *Section 2* - CT characteristics
  ○ Date and time of CT
  ○ Visible ischaemic brain lesion (Y/N)
    ▪ If Y, location of lesion:
      - Side (L/R/both/midline)
      - Location (basal ganglia, frontal lobe, parietal lobe, occipital lobe, temporal lobe, brainstem, cerebellum)
      - Cortical/ subcortical/ both
    ▪ If N, which side of brain *they expect* is affected by stroke (L/R/unknown)
  ○ Hyperdense artery (Y/N)
    ▪ If Y, location of lesion:
      - Side (L/R)
      - Named artery (Internal carotid artery, Middle cerebral artery [mainstem or sylvian branches], Anterior cerebral artery, Posterior cerebral artery, Vertebral artery, Basilar artery)
  ○ CTA imaging available (Y/N)
    ▪ If Y, is there an arterial abnormality that might account for the presenting stroke? (Y/N/Not sure)
  ○ CTP imaging available (Y/N)
    ▪ If Y, is there a perfusion defect indicating an ischaemic lesion (Y/N/not sure)
  ○ For hyperdense artery/CTA/CTP
    ▪ Whether identified abnormality helped user to identify ischaemic brain lesion on non-enhanced CT
- *Section 3* - CT Clock Tool measurements
  ○ CT attenuation within the visible acute ischaemic lesion (or if no visible acute lesion, in a region of brain the user expects to be affected by ischaemia – based on the presence of a hyperdense artery, abnormal CTA or abnormal region on CTP, or clinical examination alone).
  ○ CT attenuation within the contralateral normal brain (including equivalent proportions of grey and white matter)
- *Section 4* - Practicalities of using the CT Clock Tool
  ○ Only if an ischaemic lesion was identified:
    ▪ Finding the lesion (Grading from 1 Easy to 5 Difficult)
    ▪ Measuring the lesion (Grading from 1 Easy to 5 Difficult)
  If an ischaemic lesion was not identified:
    ▪ Estimating the location of ischaemia (Grading from 1 Easy to 5 Difficult)
    ▪ Time spent using the CT Clock Tool (1-2 minutes/3-5 minutes/More than 5 minutes)
- For electronic CT Clock Tool assessment form only:
  ○ Date-time of form submission (automatically stored)
  ○ If possible, automatic recording of time per section (especially sections 2 and 3)
  ○ To ensure completion of the form, all questions will be mandatory (individual section pages will not submit without completion) but data will be stored per page/section submission.

**No patient identifiable information will be recorded by clinicians using either the paper or the electronic CT Clock Tool assessment form.** As noted in 7.1.1, EMERGE nurses will add patient identifiable data to the paper CT Clock Tool assessment forms to ensure data are collected from the correct patient.

#### 7.1.3 Follow-up data

Collected by CI and recorded in the electronic CRF:

- Participating patient study number.
- Date and time of baseline CT acquired for immediate stroke care.
- According to expert (CI) review with access to all other data including additional imaging, whether any of the following are visible on baseline CT:
  ○ A visible ischaemic brain lesion (Y/N)
    - If Y, location and extent of lesion, using the same location method described in 7.1.2.2, but also using a more detailed and clinically-validated method^12^
  ○ A hyperattenuating artery (Y/N)
    ▪ If Y, location and extent of hyperattenuation, by named arterial segments
  ○ Old stroke lesions, chronic white matter changes, other diagnoses (Y/N for each)
- Whether CTA was acquired at baseline
  ○ Date and time of CTA
  ○ According to expert (CI) review, whether baseline CTA demonstrates a relevant arterial abnormality.
- Whether CTP was acquired at baseline
  ○ Date and time of CTP.
  ○ According to expert (CI) review, whether baseline CTP includes core and/or penumbral tissue (masked to CT Clock Tool results provided by clinicians) – scored for location using the same methods described above.
- Details of any other relevant imaging acquired during hospital stay.
- Final diagnosis – ideally confirm ischaemic stroke but might include stroke mimic caused by a structural lesion not immediately recognised.

**No patient identifiable information will be recorded at follow-up.**

#### 7.1.4 CT imaging data

Collected by the EMERGE team for secure central storage within the University of Edinburgh, Centre for Clinical Brain Sciences research PACS (Picture Archiving and Communication System):

- Pseudonymised copies (see 8.2) of all CT brain imaging associated with the hospital visit.
- Will include as minimum, a non-enhanced CT brain scan for each participant but may also include CTA and CTP acquired at baseline, and further CT and/or MRI scans acquired during follow-up (additional imaging dictated by clinical need).
- No imaging will be acquired specifically for this study.

#### 7.1.5 Data validation and missing data

Following completion of follow-up data collection, the CI will then review screening and baseline data for completeness, and will attempt to find missing data (excluding CT Clock Tool measurements, which must be completed on the day of recruitment and from which CI will be masked) either from routinely collected data sources or from the clinician involved.

### 7.2 Source Data

- All screening, baseline (except CT Clock Tool assessments) and follow-up data (see 6.1) will be transcribed from original routinely-collected healthcare data.

○ Sources for routinely-collected data include ambulance handover notes, emergency department records, stroke department records, radiology department records. The latter includes review of medical imaging on local (NHS Lothian) PACS server by CI. Sources will be assessed from baseline until follow-up review, i.e. from day 1 until day 14-21.
- Data collected during CT Clock Tool assessment (7.1.2.2) will be collected specifically for the study (not routine care) and thus represents additional, not source data.
- Routinely-collected brain imaging acquired at baseline and during the hospital admission will be copied from the NHS Lothian PACS and retained for follow-up review, see 7.1.4.

### 7.3 Case Report Forms

#### 7.3.1 Paper CRF

**Includes patient identifiable information.**

For practical purposes relating to data collection in the RIE emergency department, baseline clinical and patient data (except CT Clock Tool assessments, see 7.1.2) will be recorded in a paper CRF for each recruited participant.

The EMERGE nurses will be responsible for handling and storing paper CRFs immediately after the baseline visit and for transcribing non-identifiable data to the electronic CRF (when time allows).

#### 7.3.2 Electronic CRF

**Does not include patient identifiable information – participating patients identified using study number only, i.e. pseudonymised.**

Ultimately, all non-identifiable, non-imaging data collected for this study will be added to the electronic CRF.

The CT Clock Tool website, available at www.ctclock.net will act as the portal for data submission and will include several bespoke electronic forms securely linked to a database, see 8.3.2:

- Baseline data submission form (7.1.2.1)
- CT Clock Tool assessment form (7.1.2.2)
- Follow-up data submission form (7.1.3).

## 8 DATA MANAGEMENT

### 8.1 Patient Identifiable Data

For recruited participating patients only (but not screened, non-recruited potential participants), the following identifiable data will be collected:

- Name
- CHI number.

Name and CHI number (for patients) will be recorded on consent forms (only name for participating clinicians), on the *Consent & Subject Status Log*, on CT Clock Tool paper reminder forms, and on the paper CRF.

The *Consent & Subject Status Log* and the paper CRFs will also record each patient participant’s unique study number and will therefore act as pseudonymisation keys for electronic data. The electronic CRF and electronic CT Clock Tool assessment forms will identify participants using only their study ID number.

Although CT brain imaging will be pseudonymised following removal of participant identifiable meta data on imaging (see 8.2) replaced with the study ID number, since brain imaging often includes the face, it will still be handled as participant identifiable data.

### 8.2 Data Information Flow

Patient identifiable data will only be collected during the baseline visit and only recorded on paper documents as described in 8.1.

Non-identifiable participant data including clinical data recorded in the paper CRF and data from paper-based CT Clock Tool assessments will be transcribed to the electronic CRF by EMERGE nurses or the study CI once the baseline visit is complete.

Patient identifiable data will only be used to enable collection of the baseline and follow-up data by allowing linkage to source data (see 7.2).

Non-imaging data will be transmitted by researchers to the electronic CRF and electronic CT Clock Tool assessment forms (either directly or from previously completed paper documents) using NHS Lothian computers with simultaneous access to patient health records including imaging.

The RIE radiology department will provide digital copies of imaging data (on CD-ROMs for transfer to the University of Edinburgh) in the DICOM (Digital Imaging and Communications in Medicine) format, see 8.3.3. These DICOM data will be pseudonymised at the time of copy. Thus, patient identifiable meta data including name, date of birth, CHI number, etc., will be removed. Instead, each scan will be labelled with the participant’s unique study number. Other meta data such as date-time of scan (and other technical parameters) will be retained in the DICOM files.

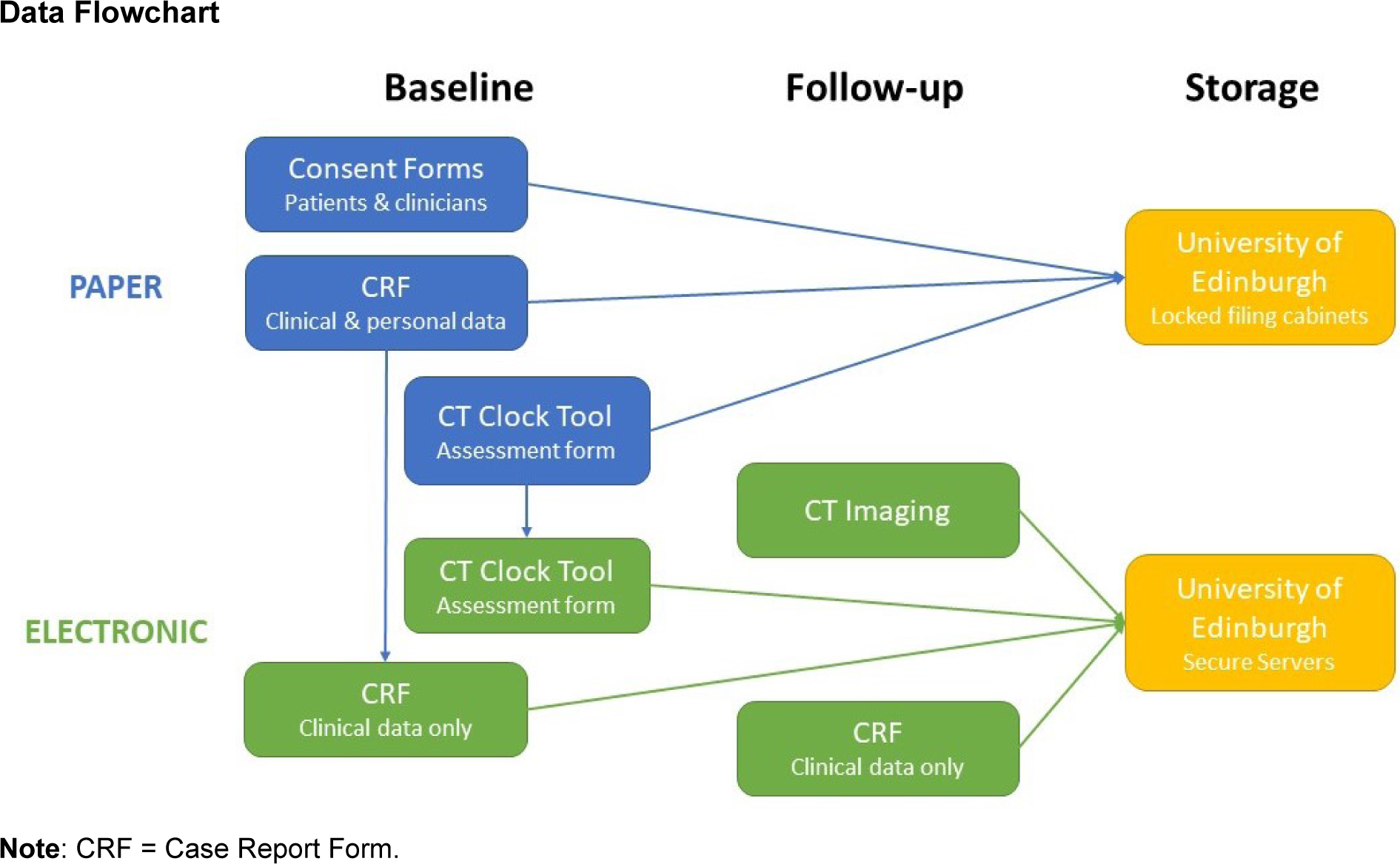

### 8.3 Data Storage

Only members of EMERGE and of the immediate research team will have access to participant identifiable data for the purposes of data collection and data validation.

Members of the research team and their designated proxies will be allowed to access pseudonymised data for the purposes of analysis.

#### 8.3.1 Data collected on paper forms (including consent)

Includes patient identifiable data.

Will be stored in locked filing cabinets in the EMERGE offices that have restricted (key card) access.

#### 8.3.2 Electronic data

Does not include patient identifiable data, cases identified using unique study IDs only.

Electronic data will be stored in a database within a secure University of Edinburgh server. The study webpage (www.ctclock.net) will be hosted on the same secure University of Edinburgh server with direct secure communication to the study database. Website and database management is only possible from within the University of Edinburgh network.

Each electronic CRF data input form will have a specific database table. Database tables will be linked using unique study IDs. Access to data input forms on www.ctclock.net will not be publicly available, but will be controlled with generic user logins defined by role:

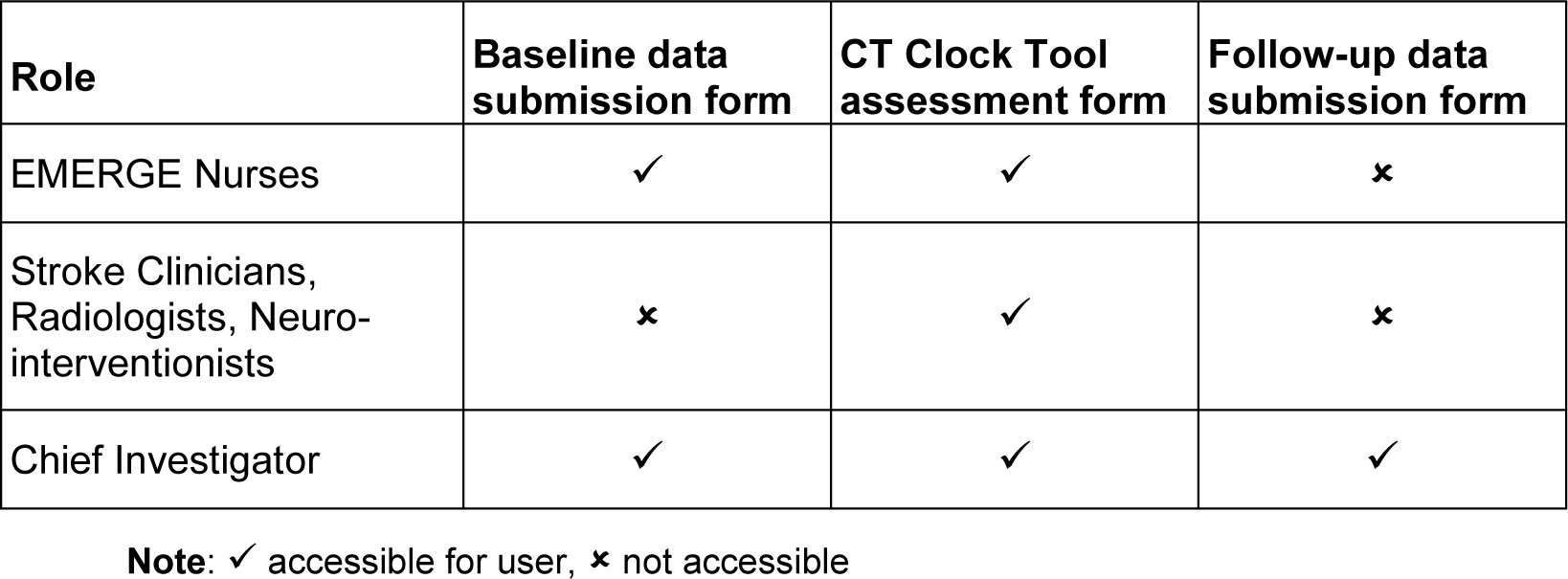

#### 8.3.3 CT imaging data

Does not include patient identifiable data.

Pseudonymised CT imaging data copied from NHS Lothians PACS will be transferred via CD-ROM to the secure research PACS server hosted within the University of Edinburgh, Centre for Clinical Brain Sciences.

We will use the Edinburgh University SMARTIS (Systematic Management, Archiving and Reviewing of Trial Images Service) service for secure and safe transfer of imaging data; http://www.ed.ac.uk/edinburgh-imaging/smartis. The EMERGE team will collect CD-ROMS for the RIE radiology department at monthly intervals and hand deliver these to SMARTIS offices, which are based within the Centre for Clinical Brain Sciences, University of Edinburgh Medical School, immediately adjacent to RIE.

Following upload of imaging to the University research PACS, CD-ROMS will be securely stored in a locked cabinet within the University of Edinburgh Centre for Clinical Brain Sciences offices.

Only the CI and SMARTIS staff will have access to the CD-ROMS. Access to the University of Edinburgh research PACS is restricted to designated users and access is only possible within the University network.

### 8.4 Data Retention

All physical data formats (paperwork including participant identifiable data and CD-ROMs including pseudonymised imaging data) will be stored for 12 months after the completion of the study (anticipated to be Sep 2025) before being destroyed.

Electronic data will be of two types, both pseudonymised:

1. Non-imaging data (e.g. age and sex of participants, details about their stroke and imaging acquired during care, measurements from CTs, clinician survey data) contained within the study database.
2. Imaging data held within PACS.

Following completion of the study, the study database will be locked, and non-imaging electronic data will be extracted from the database as spreadsheets for analysis. Spreadsheets will be saved in a widely accessible format, e.g. CSV (comma separated values) files.

Once analyses are complete, anonymised non-imaging data (i.e. pseudonymisation ID removed and spreadsheet rows randomly reordered) pertaining to that analysis will be made openly available in perpetuity, see 8.6. This will comply with the funder’s (UKRI/MRC) requirements to maximise the value of data for wider research use.

Imaging data will be stored on the University of Edinburgh research PACS indefinitely, but will not be openly shared.

Following destruction of the pseudonymisation key at 12 months, all electronic study data will become anonymised.

### 8.5 Disposal of Data

All physical data formats (paperwork including participant identifiable data and CD-ROMs including pseudonymised imaging data) will be destroyed 12 months after the study concludes. These documents will be shredded, and the waste disposed of securely using University of Edinburgh approved confidential waste streams.

The study database will be destroyed at five years after the study concludes. Database files will be enclosed in an encrypted container with a highly secure password (random string of 16 characters) before being deleted. The encrypted container password will not be retained.

### 8.6 External Transfer of Data

Data collected and generated by the study will be of two types:

1. Physical data formats (paper documents and CD-ROMs) which include participant identifiers.
2. Electronic data that does not include patient identifiers but is pseudonymised.

Physical data formats (including personal data and the pseudonymisation key) will not be transferred to any external individuals or organisations outside of the Sponsoring organisations (NHS Lothian and University of Edinburgh).

Electronic data, that are at least pseudonymised, ideally anonymised will be available for sharing, depending on format:

- Spreadsheets including anonymised non-imaging data (no included patient identifiers, pseudonymisation ID removed) will be made openly available using the University of Edinburgh’s DataShare platform, https://datashare.ed.ac.uk/.
  ○ DataShare aims to make research data access FAIR (findable, accessible, interoperable, reusable). Thus, all data has a DOI, is openly downloadable, includes a Creative Commons Attribution 4.0 International (CC-BY-4.0) license, stored in widely accessible formats, and includes relevant meta-data/plain English summaries/explanations or variable names, data types, etc.
  ○ Prior to submitting articles for publication, we will complete transfer of the shareable data used in each article to the DataShare repository. Therefore, each study output will include a DOI for the data used in that article.
- Imaging data of the brain in DICOM format may include the participant’s face and therefore may remain disclosive.
  ○ These data will not be made openly available, nor will they be routinely accessible outside the University of Edinburgh network.
  ○ However, the study CI and any of the protocol authors will be able to approve future collaborative research projects using the imaging data. Any such future projects proposing to reuse imaging data will be collaborative in nature (i.e. including members of the current research team) and for most, data will not be expected to leave the University network.
  ○ Imaging data may only be allowed to leave the University network if approved by the CI and a majority of the protocol author group.
  ○ If agreement is reached to share imaging data outside the university network, then a data sharing agreement will be required. These agreements will be drafted by the University of Edinburgh legal team, but the main responsibilities will include securely handling the data only in a pre-specified location (e.g. another University’s network), limiting access to a designated research team, acknowledging use of the data; further sharing will be precluded.
  ○ If imaging data are to be shared for commercial research or are to be shared outside the European Economic Area (EEA), then a data sharing contract will be required. These contracts will be drafted and approved by the University of Edinburgh Legal Services department and will include specific data protection contract clauses.

### 8.7 Data Controller

The University of Edinburgh and NHS Lothian are joint data controllers.

### 8.8 Data Breaches

Any data breaches will be reported to the University of Edinburgh and NHS Lothian Data Protection Officers who will onward report to the relevant authority according to the appropriate timelines if required.

## 9 STATISTICS AND DATA ANALYSIS

### 9.1 SAMPLE SIZE CALCULATION

Using methods described by Buderer,^13-15^ we aim to test whether our previous estimates for CT Clock Tool sensitivity and specificity are precise to ±10%, i.e. we will estimate the number of patients needed for two-sided 95% confidence intervals to have approximately 20% width. This level of precision would be clinically meaningful:

1. For identifying patients under 4.5 hours from ischaemic stroke onset (97% and 83%, respectively)
2. For identifying injured brain tissue that remains viable (86% and 91%, respectively)

From recent UK stroke audit data (Apr 20-March 21), the median time from stroke symptom onset to initial imaging was 4 hours and 22 minutes.^1^ Therefore, we estimate that 54% of patients are imaged within 4.5 hours, this proportion represents the prevalence used in the following calculations (we do not know the prevalence of viable brain tissue at presentation and use the same estimated prevalence since time is a surrogate for brain tissue viability):

- The number of patients required for the sensitivity and specificity of stroke onset time estimates are 21 and 118, respectively.
- The number of patients required for the sensitivity and specificity of brain tissue viability estimates are 86 and 69, respectively.

The final sample size for our prospective analysis is set at the largest estimate, therefore 118 patients in total.

There are approximately 950 presentations annually to the RIE emergency department with confirmed stroke. Of these, 86% are found to have an ischaemic aetiology while 40% present during the normal working hours covered by the EMERGE team.^16^ UK stroke audit data suggests that stroke severity (NIHSS) is assessed for 94% of patients and 50% of these have NIHSS >4 at presentation, and 69% have a known time of symptom onset.^1^ Thus, an estimated 106 patients in RIE are potential candidates for recruitment annually. With almost no requirements expected of recruited patients (except consent) and no change to their care as part of the study, recruitment rates are expected to be high, i.e. >90%. Therefore, we anticipate that recruitment of 118 patients would require up to 15 months. However, since each recruited patient can have their imaging assessed independently by up to 6 different clinicians (a consultant and a trainee stroke clinician, a consultant and a trainee radiologist, and a consultant and a trainee neurointerventionist) in parallel, fewer unique patients and less recruitment time is anticipated to reach 118 patient assessments. Thus, the absolute minimum number of patients that we need to recruit is 24 (118/5=23.6). However, assuming an average of 2-3 Clock Tool evaluations per patient we anticipate that perhaps 40-60 patients are required. We will recruit for up to 12 months to allow for incomplete stroke overlap between stroke clinicians and radiologists assessing cases; a recruitment time of 12 months is expected to be sufficient.

### 9.2 PROPOSED ANALYSES

#### 9.2.1 Primary objective

To evaluate the feasibility of front-line clinicians using the CT Clock Tool during routine care, we will assess:

- How often CT Clock Tool method is successfully used
- Ease of use
  ○ Finding and measuring a lesion
  ○ Calculating attenuation ratio
  ○ Impact of other imaging features
- Time taken.

We will present feasibility data as proportion of:

- Cases where a front-line clinician successfully uses the CT Clock Tool method (regardless of result)
- Front-line clinicians correctly identifying acute ischaemic lesions on baseline non-enhanced CT alone (i.e. where concurrent CT perfusion is not available) compared to an expert using all available data including follow-up imaging.
- Baseline non-enhanced CT brain scans that include other findings that might confound results (e.g. old stroke lesions, chronic white matter changes, secondary diagnoses)
- Cases where the time needed to use the CT Clock Tool was less than 5 minutes.

##### 9.2.1.1 Success criteria for primary objective

1. Where an ischaemic lesion is identified, we expect that >50% of clinicians will report that measuring these lesions was ‘easy’.
2. For all users, we expect that >50% of clinicians will report that using the CT Clock Tool was quick or had minimal delay (i.e. less than 5 minutes). If it is possible to automatically record the time users spend completing the electronic CRF, we expect that >50% of clinicians will spend <5 minutes using the CT Clock Tool (i.e. finding & measuring lesions and inputting these details to the form, in other words completing sections 2-3 of electronic CRF, see 7.1.2.2).

#### 9.2.2 Secondary objectives

To evaluate CT Clock Tool results and their potential impact on clinical care, we will derive estimates for time elapsed (in minutes) from stroke onset time to CT using the CT attenuation measurements provided by clinicians, and we will assess:

- Whether stroke onset time estimates provided by front-line clinicians using the CT Clock Tool are similar to actual elapsed time (where stroke onset time is known)

○ Difference in minutes between true elapsed time and CT Clock Tool time estimates
○ Absolute error of time estimates
○ Proportion of cases where estimates over or under call elapsed time
○ Correlation between error and true elapsed time.
- Diagnostic accuracy of CT Clock Tool time estimates for determining treatment eligibility relative to standard thrombolysis and thrombectomy time limits (4.5 hours and 6 hours, respectively)

○ Sensitivity, specificity, positive and negative predictive values
○ Receiver operating characteristic (ROC) curve analysis.
○ In whole group and in subgroups:

▪ Visible lesion vs estimated lesion location
▪ Milder (NIHSS less than 6, 9, or 12) vs more severe stroke
- Whether CT Clock Tool estimates of brain tissue viability predict equivalent CT perfusion findings

○ Where concurrently acquired CT perfusion is available, proportion where front-line clinician derived attenuation ratio correctly predicts viable/reversible brain tissue injury (penumbra) defined using CTP.
- User experience of the CT Clock Tool

○ Mean score for ease of finding/estimating the location of lesions, and for measuring lesions.
○ Mean time spent using the CT Clock Tool method.
- Inter-rater agreement between the CT Clock Tool results provided by different clinicians for the same CT scan

○ Agreement for brain tissue measurements, attenuation ratio, time and tissue viability estimates derived from these attenuation ratios.

##### 9.2.2.1 Success criteria for secondary objectives

1. Sensitivity and specificity >70% at identifying patients who are within 4.5 hours of stroke symptom onset.
2. An attenuation ratio >0.87 correctly predicts the presence of potentially reversible tissue injury (penumbra) on perfusion imaging on >70% of patient assessments.

#### 9.2.3 Statistics

We will

- Use descriptive statistics to define the cohort, and predominantly use proportions to assess objectives
- Use Bland-Altman methods^17^ to compare estimated and true elapsed time
- Use univariate statistics to assess correlation and to compare results between different types of clinicians using the CT Clock Tool

○ Radiologists vs stroke clinicians
○ Consultants vs trainees
- Derive true and false positive and negative cases (based on CT Clock Tool results being less than thrombolysis and thrombectomy time thresholds) and we will calculate diagnostic accuracy statistics as standard.
- Use Krippendorff’s Alpha^18^ to assess inter-rater agreement.
- Preferentially report 95% confidence intervals but will also consider a p-value <0.05 significant
- Not impute but will report missing data, including any withdrawals.

#### 9.2.4 Interim analysis

To allow for timely planning of a potential follow-on multicentre evaluation of the CT Clock Tool for patients with an unknown time of stroke symptom onset, we will conduct an interim analysis at the midpoint of the study.

Midpoint is defined as having acquired 50% of proposed CT Clock Tool evaluations (i.e. 59), or reaching six-months from the study start date, whichever is sooner.

The interim analysis will define the partial cohort, assess the primary objective (ease of use and time taken), and assess agreement between CT Clock Tool estimates of elapsed time and actual elapsed time.

## 10 OVERSIGHT ARRANGEMENTS

### 10.1 INSPECTION OF RECORDS

Study investigators will permit study related monitoring and audits on behalf of the Sponsor, REC review, and regulatory inspection(s). In the event of audit or monitoring, the CI agrees to allow the representatives of the Sponsor direct access to all study records and source documentation. In the event of regulatory inspection, the CI agrees to allow inspectors direct access to all study records and source documentation.

### 10.2 STUDY MONITORING AND AUDIT

The ACCORD Sponsor Representative will assess the study to determine if a study specific risk assessment is required.

If required, a study specific risk assessment will be performed by representatives of the Sponsor(s), ACCORD monitors and the QA group, in accordance with ACCORD governance and sponsorship SOPs. Input will be sought from the CI or designee. The outcomes of the risk assessment will form the basis of the monitoring plans and audit plans.

If considered necessary, ACCORD clinical trial monitors, or designees, will perform monitoring activities in accordance with the study monitoring plan. This will involve on-site visits and remote monitoring activities as necessary. ACCORD QA personnel, or designees, will perform study audits in accordance with the study audit plan. This will involve study management audits and facility (including 3^rd^ parties) audits as necessary.

## 11 GOOD CLINICAL PRACTICE

### 11.1 ETHICAL CONDUCT

The study will be conducted in accordance with the principles of the International Conference on Harmonisation Tripartite Guideline for Good Clinical Practice (ICH GCP).

Before the study can commence, all necessary approvals will be obtained and any conditions of approvals will be met.

### 11.2 CHIEF INVESTIGATOR RESPONSIBILITIES

The CI is responsible for the overall conduct of the study at the site and compliance with the protocol and any protocol amendments. In accordance with the principles of ICH GCP, the following areas listed in this section are also the responsibility of the CI. Responsibilities may be delegated to an appropriate member of study site staff.

Delegated tasks must be documented on a Delegation Log and signed by all those named on the list prior to undertaking applicable study-related procedures.

#### 11.2.1 Informed Consent

The CI is responsible for ensuring informed consent is obtained before any study specific procedures are carried out. The decision of a participant to participate in clinical research is voluntary and should be based on a clear understanding of what is involved.

Participants must receive adequate oral and written information – appropriate Participant Information and Informed Consent Forms will be provided. The oral explanation to the participant will be performed by the CI or qualified delegated person, and must cover all the elements specified in the Participant Information Sheet and Consent Form.

The participant must be given every opportunity to clarify any points they do not understand and, if necessary, ask for more information. The participant must be given sufficient time to consider the information provided. It should be emphasised that the participant may withdraw their consent to participate at any time without loss of benefits to which they otherwise would be entitled.

Patient participants will be informed and agree to their medical records being inspected by regulatory authorities and representatives of the Sponsor(s).

The CI or delegated member of the study team and the participant will sign and date the Informed Consent Form(s) to confirm that consent has been obtained. The original will be signed in the Investigator Site File (ISF). The participant will receive a copy of the signed consent form and for patient participants (not clinician participants) a copy will be filed in the participant’s medical notes.

#### 11.2.2 Study Site Staff

The CI is responsible for ensuring that all staff assisting with the study are adequately informed about the protocol and their study related duties.

#### 11.2.3 Data Recording

The CI is responsible for the quality of the data recorded in the CRF.

#### 11.2.4 GCP Training

For non-CTIMP (i.e. non-drug) studies all researchers are encouraged to undertake GCP training in order to understand the principles of GCP. This is not a mandatory requirement unless deemed so by the Sponsor. GCP training status for all investigators should be indicated in their respective CVs.

#### 11.2.5 Data Protection Training

All University of Edinburgh employed researchers and study staff will complete the Data Protection Training through Learn.

NHS Lothian employed researchers and study staff will comply with NHS Lothian mandatory Information Governance Data Protection training through LearnPro.

Non-NHS Lothian staff that have access to NHS Lothian systems will familiarise themselves and abide by all NHS Lothian IT policies, as well as employer policies.

#### 11.2.6 Information Security Training

All University of Edinburgh employed researchers, students and study staff will complete the Information Security Essentials modules through Learn and will have read the minimum and required reading setting out ground rules to be complied with.

NHS Lothian employed researchers and study staff will comply with NHS Lothian mandatory Information Governance IT Security training through LearnPro.

Non-NHS Lothian staff that have access to NHS Lothian systems will familiarise themselves and abide by all NHS Lothian IT policies, as well as employer policies.

#### 11.2.7 Confidentiality

All evaluation forms, reports, and other records must be identified in a manner designed to maintain participant confidentiality. All records must be kept in a secure storage area with limited access. Clinical information will not be released without the written permission of the participant. The CI and study site staff involved with this study may not disclose or use for any purpose other than performance of the study, any data, record, or other unpublished information, which is confidential or identifiable, and has been disclosed to those individuals for the purpose of the study. Prior written agreement from the Sponsor or its designee must be obtained for the disclosure of any said confidential information to other parties.

#### 11.2.8 Data Protection

All Investigators and study site staff involved with this study must comply with the requirements of the appropriate data protection legislation (including the General Data Protection Regulation and Data Protection Act) with regard to the collection, storage, processing and disclosure of personal information.

Computers used to collate the data will have limited access measures via user names and passwords.

Published results will not contain any personal data that could allow identification of individual participants.

## 12 STUDY CONDUCT RESPONSIBILITIES

### 12.1 PROTOCOL AMENDMENTS

Any changes in research activity, except those necessary to remove an apparent, immediate hazard to the participant in the case of an urgent safety measure, must be reviewed and approved by the CI.

Proposed amendments will be submitted to the Sponsor for classification, review and authorisation.

Amendments to the protocol must be submitted in writing to the appropriate REC and local R&D for approval prior to implementation and prior to participants being enrolled into the amended protocol.

### 12.2 MANAGEMENT OF PROTOCOL NON COMPLIANCE

#### 12.2.1 Protocol Waivers

Prospective protocol deviations, i.e. protocol waivers, will not be approved by the Sponsors and therefore will not be implemented, except where necessary to eliminate an immediate hazard to study participants. If this necessitates a subsequent protocol amendment, this should be submitted to the REC and local R&D for review and approval if appropriate.

#### 12.2.2 Management of Deviations and Violations

Deviations and violations are non-compliance events discovered after the event has occurred. Protocol deviations will be recorded in a protocol deviation log and logs will be submitted to the Sponsors every 3 months. Each protocol violation will be reported to the Sponsor within 3 days of becoming aware of the violation.

Deviation logs/violation forms will be transmitted via email to. Only forms in a pdf format will be accepted by ACCORD via email. Forms may also be submitted by hand to the office. Where missing information has not been sent to ACCORD after an initial report, ACCORD will contact the CI and request the missing information. The CI must respond to these requests in a timely manner.

### 12.3 SERIOUS BREACH REQUIREMENTS

A serious breach is a breach which is likely to effect to a significant degree:

(a) the safety or physical or mental integrity of the participants of the study; or

(b) the scientific value of the study.

If a potential serious breach is identified by the CI or delegates, the Sponsor(s) must be notified within 24 hours. It is the responsibility of the Sponsor(s) to assess the impact of the breach on the scientific value of the study, to determine whether the incident constitutes a serious breach and report to research ethics committees as necessary.

### 12.4 STUDY RECORD RETENTION

All study documentation will be kept for a minimum of 3 years from the protocol defined end of study point. When the minimum retention period has elapsed, study documentation will be destroyed with permission from the Sponsor.

### 12.5 END OF STUDY

The end of study is defined as the last participant’s last visit.

The CI and/or the Sponsor(s) have the right at any time to terminate the study for clinical or administrative reasons.

The end of the study will be reported to the REC, and R&D Office(s) and Sponsor(s) within 90 days, or 15 days if the study is terminated prematurely. The CI will inform participants of the premature study closure and ensure that the appropriate follow up is arranged for all participants involved. End of study notification will be reported to the Sponsor(s) via email to.

A summary report of the study will be provided to the REC within 1 year of the end of the study.

### 12.6 INSURANCE AND INDEMNITY

The Sponsor(s) are responsible for ensuring proper provision has been made for insurance or indemnity to cover their liability and the liability of the CI and staff.

The following arrangements are in place to fulfil the Sponsor(s)’ responsibilities:

- The Protocol has been designed by the CI and researchers employed by the University and collaborators. The University has insurance in place (which includes no-fault compensation) for negligent harm caused by poor protocol design by the CI and researchers employed by the University.
- Sites participating in the study will be liable for clinical negligence and other negligent harm to individuals taking part in the study and covered by the duty of care owed to them by the sites concerned. The Sponsor(s) require individual sites participating in the study to arrange for their own insurance or indemnity in respect of these liabilities.
- Sites which are part of the United Kingdom’s National Health Service will have the benefit of NHS Indemnity.

## 13 AUTHORSHIP POLICY

Ownership of the data arising from this study resides with the study team.

## Data Availability

Following study completion data will be available online at https://datashare.ed.ac.uk/

## LIST OF ABBREVIATIONS

ACCORD: Academic and Clinical Central Office for Research & Development - Joint office for The University of Edinburgh and Lothian Health Board
CRF: Case Report Form
CSV: Comma Separated Values
CHI: Community Health Index
CI: Chief Investigator
CIS: Clinician Information Sheet
CT: Computed Tomography imaging
CTA: CT Angiography imaging
CTP: CT Perfusion imaging
DICOM: Digital Imaging and Communications in Medicine
EMERGE: Emergency Medicine Research Group of Edinburgh
FIS: Family Information Sheet
GCP: Good Clinical Practice
NHS: National Health Service
NIHSS: National Institutes of Health Stroke Scale
MRC: Medical Research Council
MRI: Magnetic Resonance Imaging
OCSP: Oxford Community Stroke Project Classification
PACS: Picture Archiving and Communication System
PIS: Patient Information Sheet
QA: Quality Assurance
REC: Research Ethics Committee
RIE: Royal Infirmary of Edinburgh
SMARTIS: Systematic Management, Archiving and Reviewing of Trial Images Service
SOP: Standard Operating Procedure
UKRI: UK Research and Innovation

## REFERENCES

1. Sentinel Stroke National Audit Programme (SSNAP): 9th Annual Report (Apr 2021 to March 2022). Retrieved 12th January, 2023, from https://www.strokeaudit.org/results/Clinical-audit/National-Results.aspx

2. Albers GW, Marks MP, Kemp S, Christensen S, Tsai JP, et al. Thrombectomy for Stroke at 6 to 16 Hours with Selection by Perfusion Imaging. N Engl J Med. 2018;378:708–718. 10.1056/NEJMoa1713973

3. Nogueira RG, Jadhav AP, Haussen DC, Bonafe A, Budzik RF, et al. Thrombectomy 6 to 24 Hours after Stroke with a Mismatch between Deficit and Infarct. N Engl J Med. 2018;378:11–21. 10.1056/NEJMoa1706442

4. Thomalla G, Simonsen CZ, Boutitie F, Andersen G, Berthezene Y, et al. MRI-Guided Thrombolysis for Stroke with Unknown Time of Onset. N Engl J Med. 2018;379:611–622. 10.1056/NEJMoa1804355

5. Ma H, Campbell BCV, Parsons MW, Churilov L, Levi CR, et al. Thrombolysis Guided by Perfusion Imaging up to 9 Hours after Onset of Stroke. N Engl J Med. 2019;380:1795–1803. 10.1056/NEJMoa1813046

6. Mair G, Alzahrani A, Lindley RI, Sandercock PAG, Wardlaw JM. Feasibility and diagnostic accuracy of using brain attenuation changes on CT to estimate time of ischemic stroke onset. Neuroradiology. 2021;63:869–878. 10.1007/s00234-020-02591-w

7. Alzahrani A, Zhang X, Albukhari A, Wardlaw JM, Mair G. Assessing Brain Tissue Viability on Nonenhanced Computed Tomography After Ischemic Stroke. Stroke. 2023. 10.1161/STROKEAHA.122.041241

8. Mair G, Wardlaw JM. Normal Appearing Ischaemic Brain Tissue on CT and Outcome After Intravenous Alteplase. Frontiers in Radiology. 2022;2. 10.3389/fradi.2022.902165

9. Urrutia VC, Faigle R, Zeiler SR, Marsh EB, Bahouth M, et al. Safety of intravenous alteplase within 4.5 hours for patients awakening with stroke symptoms. PLoS One. 2018;13:e0197714. 10.1371/journal.pone.0197714

10. Sykora M, Kellert L, Michel P, Eskandari A, Feil K, et al. Thrombolysis in Stroke With Unknown Onset Based on Non-Contrast Computerized Tomography (TRUST CT). J Am Heart Assoc. 2020;9:e014265. 10.1161/JAHA.119.014265

11. Roaldsen MB, Eltoft A, Wilsgaard T, Christensen H, Engelter ST, et al. Safety and efficacy of tenecteplase in patients with wake-up stroke assessed by non-contrast CT (TWIST): a multicentre, open-label, randomised controlled trial. Lancet Neurol. 2023;22:117–126. 10.1016/S1474-4422(22)00484-7

12. Wardlaw JM, Sellar RJ. A simple practical classification of cerebral infarcts on CT and its interobserver reliability. AJNR Am J Neuroradiol. 1994;15:1933–1939.

13. Buderer NM. Statistical methodology: I. Incorporating the prevalence of disease into the sample size calculation for sensitivity and specificity. Acad Emerg Med. 1996;3:895–900. 10.1111/j.1553-2712.1996.tb03538.x

14. Arifin WN. Sample Size Calculator. Retrieved 21st April 2022, from https://wnarifin.github.io/ssc/sssnsp.html

15. Leeflang MMG, Allerberger F. Sample size calculations for diagnostic studies. Clin Microbiol Infect. 2019;25:777–778. 10.1016/j.cmi.2019.04.011

16. Scottish Stroke Care Audit (SSCA). Scottish stroke improvement programme: 2022 national report. Retrieved 12th January, 2023, from https://www.strokeaudit.scot.nhs.uk/index.html

17. Bland JM, Altman DG. Statistical methods for assessing agreement between two methods of clinical measurement. Lancet. 1986;1:307–310.

18. Hayes AF, Krippendorff K. Answering the Call for a Standard Reliability Measure for Coding Data. Communication Methods and Measures. 2007;1:77–89.

